# Pooling RT-PCR or NGS samples has the potential to cost-effectively generate estimates of COVID-19 prevalence in resource limited environments

**DOI:** 10.1101/2020.04.03.20051995

**Authors:** Krishna R Narayanan, Isabel Frost, Anoosheh Heidarzadeh, Katie K Tseng, Sayantan Banerjee, Jacob John, Ramanan Laxminarayan

**Affiliations:** Department of Electrical and Computer Engineering, Texas A&M University, College Station, TX 77840, USA; Center for Disease Dynamics, Economics & Policy, Silver Spring, MD 20910, USA; Imperial College London, UK; World Health Organization, South East Asia Regional Office, New Delhi, India; Dept of Community Health, Christian Medical College, Vellore 632002, India; Department of Global Health, University of Washington, Seattle, WA 98104, USA; Princeton Environment Institute, Princeton University, Princeton, NJ 08544, USA

## Abstract

**Background:** COVID-19 originated in China and has quickly spread worldwide causing a pandemic. Countries need rapid data on the prevalence of the virus in communities to enable rapid containment. However, the equipment, human and laboratory resources required for conducting individual RT-PCR is prohibitive. One technique to reduce the number of tests required is the pooling of samples for analysis by RT-PCR prior to testing.

**Methods:** We conducted a mathematical analysis of pooling strategies for infection rate classification using group testing and for the identification of individuals by testing pooled clusters of samples.

**Findings:** On the basis of the proposed pooled testing strategy we calculate the probability of false alarm, the probability of detection, and the average number of tests required as a function of the pool size. We find that when the sample size is 256, using a maximum pool size of 64, with only 7.3 tests on average, we can distinguish between prevalences of 1% and 5% with a probability of detection of 95% and probability of false alarm of 4%.

**Interpretation:** The pooling of RT-PCR samples is a cost-effective technique for providing much-needed course-grained data on the prevalence of COVID-19. This is a powerful tool in providing countries with information that can facilitate a response to the pandemic that is evidence-based and saves the most lives possible with the resources available.

**Funding:** Bill & Melinda Gates Foundation

**Authors contributions:** RL and KRN conceived the study. IF, KT, KRN, SB and RL all contributed to the writing of the manuscript and AH and JJ provided comments. KRN and AH conducted the analysis and designed the figures.

**Research in context:** *Evidence before this study:* The pooling of RT-PCR samples has been shown to be effective in screening for HIV, Chlamydia, Malaria, and influenza, among other pathogens in human health. In agriculture, this method has been used to assess the prevalence of many pathogens, including *Dichelobacter nodosus*, which causes footrot in sheep, postweaning multisystemic wasting syndrome, and antibiotic resistance in swine feces, in addition to the identification of coronaviruses in multiple bat species. In relation to the current pandemic, researchers in multiple countries have begun to employ this technique to investigate samples for COVID-19.

*Added value of this study:* Given recent interest in this topic, this study provides a mathematical analysis of infection rate classification using group testing and calculates the probability of false alarm, the probability of detection, and the average number of tests required as a function of the pool size. In addition the identification of individuals by pooled cluster testing is evaluated.

*Implications of all the available evidence:* This research suggests the pooling of RT-PCR samples for testing can provide a cheap and effective way of gathering much needed data on the prevalence of COVID-19 and identifying infected individuals in the community, where it may be infeasible to carry out a high number of tests. This will enable countries to use stretched resources in the most appropriate way possible, providing valuable data that can inform an evidence-based response to the pandemic.

---

The novel coronavirus, COVID-19, first detected in Wuhan, China, in late 2019, has become a rapidly emerging threat that has spread throughout the world. With a crude case fatality rate of 5.2%,^1^ COVID-19 poses the highest risk for serious disease and death in the over 65 year-old population.^2^ Current early stage models predict that the virus will continue to spread in the coming months and many health systems will be overwhelmed.^3^ However, these models require data on a large scale to inform an evidence-based reaction to COVID-19 that uses limited resources as effectively as possible on a rapid timescale.

Tracking the spread of COVID-19 is essential to mounting an effective public health response, understanding the current impact of the virus, and ensuring that health systems are prepared. The most accurate way of identifying COVID-19 is through the use of reverse transcription polymerase chain reaction or RT-PCR on samples.^4^ However, facilities for RT-PCR are limited, even in high-income countries, and further, test costs are high both in terms of consumables and trained technicians. Delays in testing have slowed the public health response in Europe and the United States, and challenges to testing in low- and middle-income countries are even greater.

Many countries in which transmission of COVID-19 is occurring in the community are in the acceleration phase and data describing community prevalence of the virus is much needed.

Given limitations in public health capacity, it is essential to identify high-risk clusters of infected individuals while in the stage of community containment. In the absence of sufficient RT-PCR capacity, we propose an alternative that may provide this essential data in a way that is course-grained but at lower cost using the pooling of samples from multiple patients and performing RT-PCR on this combined sample.^5–8^ If the test is negative, this implies that no one in the pool is infected and if the test is positive, it may be concluded that at least one person in the pool is infected. Pooling and group testing can be very effective in reducing the number of tests required to both identify infected people in a population and to estimate the infection rate in a population. Another benefit of this method is that it is likely to reduce testing time as fewer samples need to be prepped and put through the RT-PCR process. This technique has been shown to be effective in screening for bacteria, parasites and viruses in human health, including HIV,^5^ Chlamydia,^9^ Malaria,^9^ and influenza.^7^ In the agricultural sector, where cost-effectiveness is essential to efficient food production, the epidemiology of various pathogens, including *Dichelobacter nodosus*, which causes footrot in sheep,^10^ postweaning multisystemic wasting syndrome,^11^ and antibiotic resistance in swine feces^12^ has been measured using this method. Pooled sampling and testing via PCR has also been used to successfully identify coronaviruses in multiple bat species.^13^

## Infection rate classification using group testing

Group testing can be very beneficial in reducing the number of tests required to assess whether the infection rate in a population is low or high. We conducted a mathematical analysis of pooled sample testing in which a group of *N* people is randomly selected from the region and split into *L* subpools, each of size *N*/*L*. A subpool is infected if at least one person within it is carrying the virus. Then, we perform group testing using binary splitting at the subpool level to determine the number of subpools that are infected. We set a threshold *V*, whereby if *V*or fewer subpools are infected, we accept the low rate of infection, and if more than *V* subpools are infected, we accept the high rate of infection.

For example, let’s take a low rate of 1%and a high rate of 5%. We choose *N*=64 people from a region and form *L* =4 subpools labeled S1, S2, S3, and S4.This number has been selected as the maximum number of samples per pool as previous work from other groups suggests it is possible to detect a single positive sample in a pooled sample of 64.^16^ Finally, we choose our threshold *V*to be 1. Our group testing procedure is shown in the flowchart in **Error! Reference source not found**..

The initial test, T_1_, is performed by pooling the samples from all four subpools, S1, S2, S3, and S4. If the result of T_1_ is negative, we accept the low rate of infection. If the result of T_1_ is positive, we perform two more tests, T_2_ (containing samples S1 and S2) and T_3_ (containing samples S3 and S4). If the results of both T_2_ and T_3_ are positive, we accept the high rate of infection. If the result of only one test, T_1_ or T_2,_ is positive, then we perform two more tests by breaking that pool into two smaller subpools. For example, if the result of T_2_ is positive, then we perform two more tests, T_4_ on sample S3 and T_5_ on sample S4. If the result of only one test, T_4_ or T_5,_ is positive, then we accept the low infection rate. If the results of both tests are positive, we accept the high infection rate.

We can analyze the probability of false alarm, the probability of detection, and the average number of tests required. The distribution of the number of infected subpools, the chosen hypothesis, and the number of tests required in each case are shown in Table 1 for four subpools (*L* =4).

**Table 1.** shows the number of tests required to detect a given infection rate.

| Number of infected subpools | Infection rate | Number of tests required ($T$) |
| --- | --- | --- |
| 0 | 1% (low) | 1 |
| 1 | 1% (low) | 5 |
| 2 | 5% (high) | 3 or 5 |
| 3 | 5% (high) | 3 |
| 4 | 5% (high) | 3 |

Table 1 describes the number of tests required depending how many subpools are infected. When there are two infected subpools three or five tests are required. For example, if samples S1 and S2 are infected, five tests are required; whereas if S1 and S3 are infected, only 3 tests are required. When the number of people being tested, *N*, is large, the result of the initial test of all pooled samples, T_1_, will be positive with high probability whether the infection rate is low or high. In this case, testing the all pooled samples together will not be very informative and this test can be skipped to directly start with tests at the next level, namely T_2_ and T_3_. This strategy will reduce the number of tests without affecting the probability of false alarms or the probability of detection.

An important difference between the strategy suggested here and conventional binary splitting is that here we perform binary splitting only until we are able to ascertain if the number of infected subpools is greater than the threshold, *V*, or not; we do not necessarily infer all the infected subpools. Hence fewer tests are required on average compared to conventional binary splitting.

The probability of false alarm, the probability of detection, and the average number of tests required are shown in Figure 1 as a function of the sample size, with samples divided into eight pools and a threshold of 4 (i.e. *L* =8, *V*=4). The maximum pool size used for the tests is 64. The probability of detection increases with sample size, however the probability of false alarm is also increased. Figure 2 shows the tradeoff between the probability of detection and the probability of false alarm for two sets of parameters.

**Figure 1.**
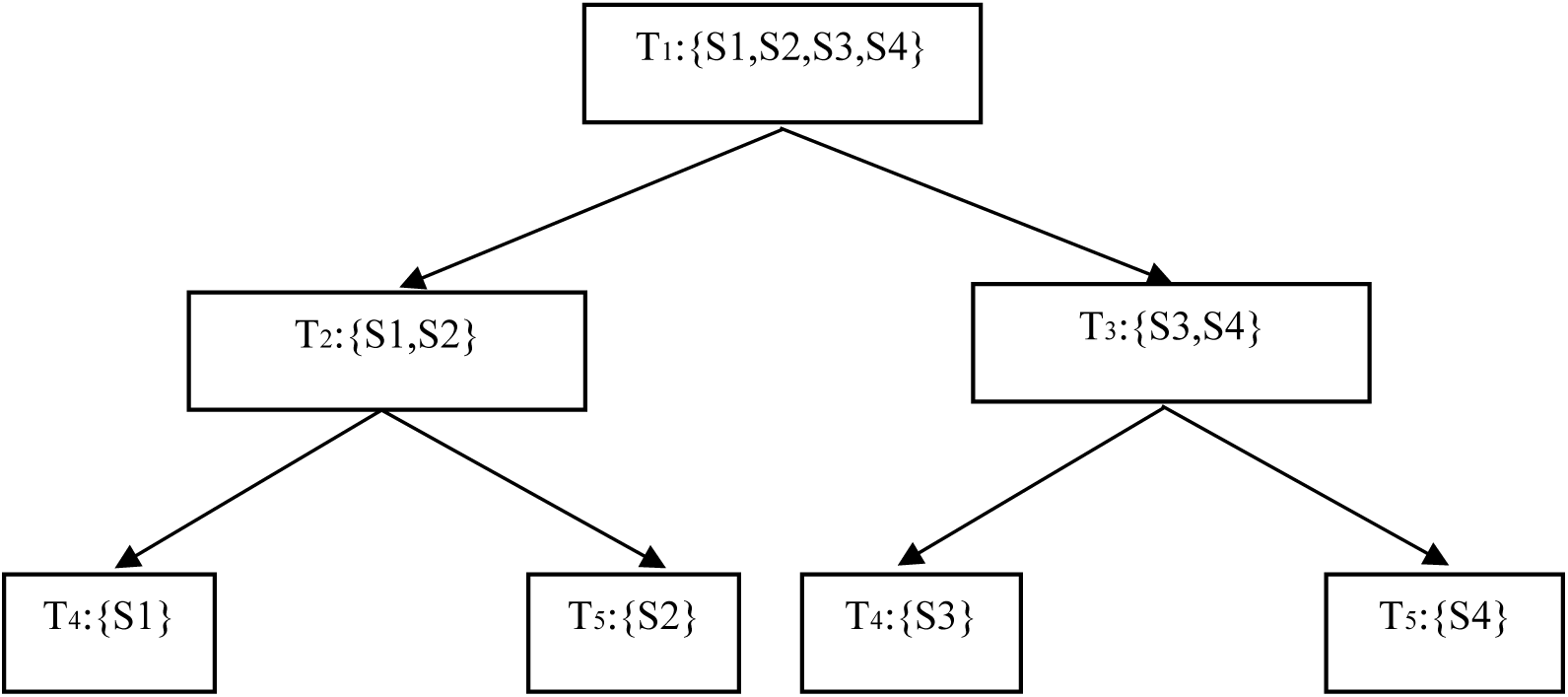
Flowchart showing the binary splitting procedure.

**Figure 1.**
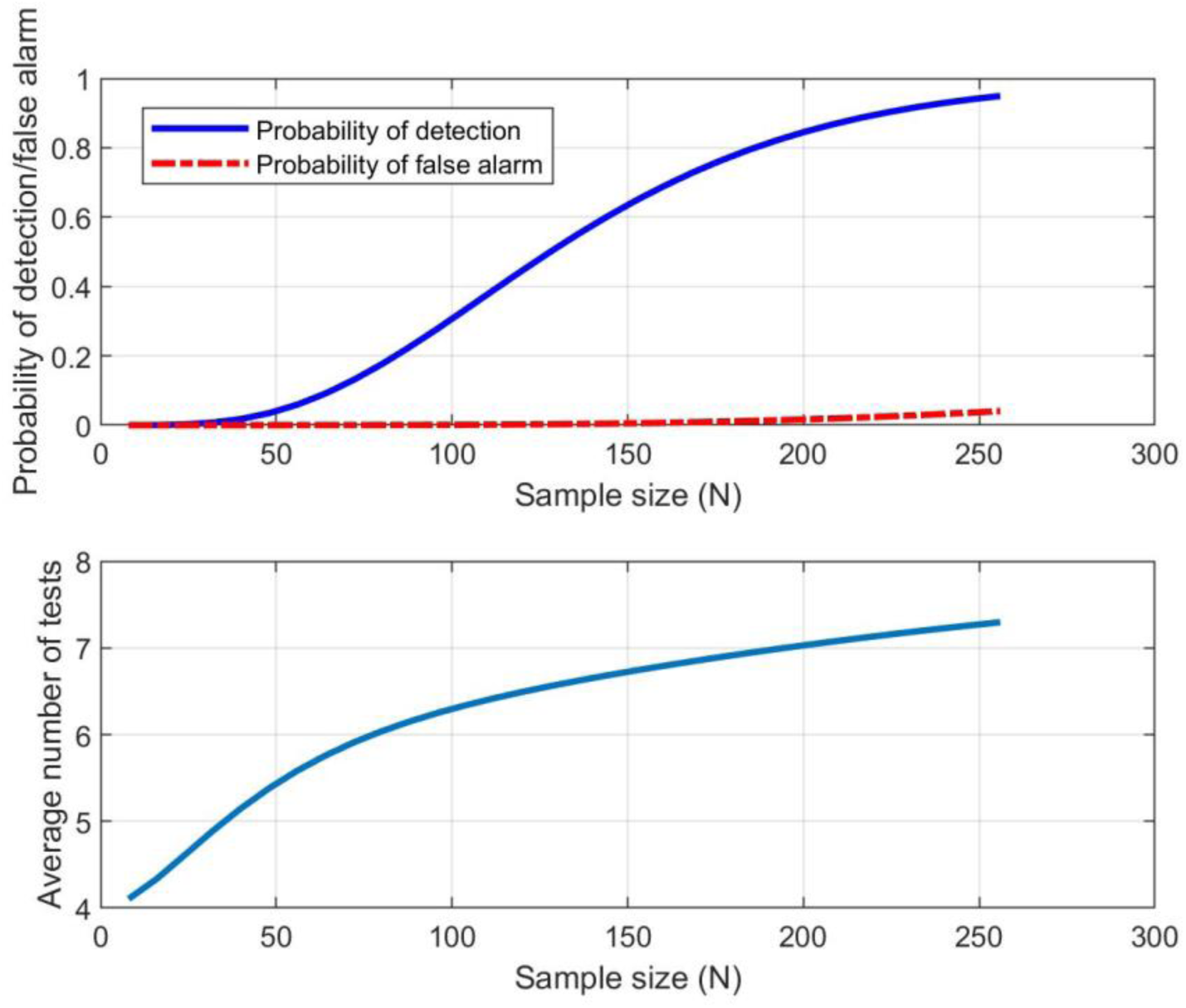
(Top) The probability of detection and false alarm as a function of N, and (Bottom) the average number of tests required as a function of N.

**Figure 2.**
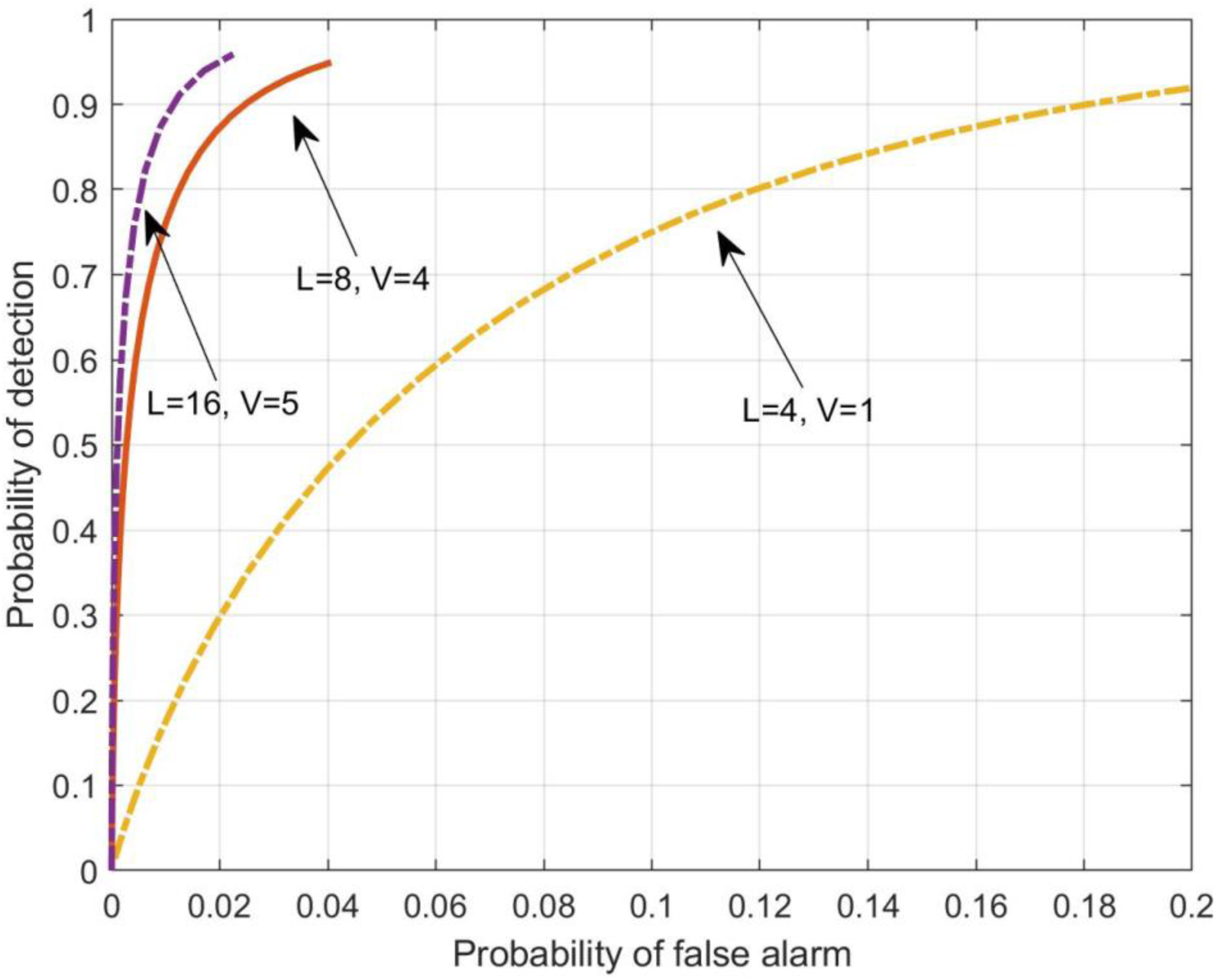
The region of convergence curve depicting the probability of detection versus the probability of false alarm for three sets of parameters.

This method of pooling samples has the potential to dramatically reduce the number of tests to be performed and this is likely to be associated with a substantial reduction in costs. In addition, as the sample size grows, it becomes easier to distinguish between high and low prevalence rates. When the sample size is of the order of 1000s, the probability of false alarm and probability of detection are greatly reduced. For example, given 256 samples and a number of samples per pool of 64, just 7.3 tests are needed on average to differentiate 1% prevalence from 5% prevalence with a probability of detection of 95% and a probability of false alarm of 4%. Further examples are provided in Table 2.

**Table 2.** Average number of tests to be performed to assess whether COVID-19 prevalence is 1% or 5% for a given number of samples.

| Number of samples | Maximum pool size | Average number of tests | Probability of detection | Probability of false alarm |
| --- | --- | --- | --- | --- |
| 200 | 50 | 5.9 | 95.7% | 7% |
| 208 | 32 | 9.3 | 96% | 2% |
| 256 | 64 | 7.3 | 95% | 4% |

### Identification of infected individuals by pooled sample testing

Next we turn to the use of pooled testing to identify infected individuals. There are many options for constructing sets of pools.^14^ One of them is the use of a binary search tree,^15^ particularly when the prevalence of COVID-19 is low. Samples may be collected in a cluster of say 32 individuals from households in close vicinity, packed and labelled individually and mapped locally by healthcare workers. In the laboratory, each cluster of samples is divided into two groups. The total number of samples in each group should typically be the largest power of 2 that gives the lowest acceptable sensitivity for the given test – either RT-PCR or next-generation sequencing (NGS), which is reported to have higher sensitivity for COVID-19.

To use the example case of a cluster of 32 samples, the group of samples pooled together shall be 16 each. Given the COVID-19 outbreak in a locality is spatially clustered, each batch or cluster of samples should consist of individual samples from houses in close vicinity to each other. This can be arranged by mapping the locality or the urban slum, plotting the households, and making sure that the samples are taken from contiguous clumps of households. After thoroughly mixing the samples individually, defined quantities of 16 samples may be pooled through a common appropriate membrane filter, and the filtrate reconstituted and tested by RT-PCR or NGS. If the pooled sample tests negative, none of the individual samples in that batch are infected. If the pooled sample tests positive, it is equally divided into two batches of half the original size, and these are tested again in the same manner.

Considering the *maximum* kit requirement situation, if there is nobody infected in the population cluster of 32 individuals, it will be detected in a single test. If there is a single infected individual among the cluster of 32, then the number of tests required will vary between 5 and 10 assuming a maximum pool size of 16. In *minimum* kit requirement situations, the kit requirements will be even smaller.

## Discussion

For evolving and novel outbreaks of infectious diseases like COVID-19, widespread individual tests are not only costly but also time consuming. Pooling of samples can help accelerate the surveillance for COVID-19 identification in a community or group of people living together. If found positive, tracing back to the individual(s) can be done by a pooled sampling exercise. In some countries this technique is already being used to make COVID-19 testing more cost-effective. In Israel, researchers at Technion – Israel Institute of Technology and Rambam Health Care Campus successfully identified a positive carrier from a pooled analysis of 64 samples.^16^ In the US, the Nebraska Public Health Lab pooled 60 samples obtained from across the state.^17^ These samples were tested in groups of five, and two groups were found to be positive. The samples from each positive group were then tested individually to identify positive carriers and the use of this technique halved the consumption of reagents. Blood banks commonly use pooled testing, combining 4 to 24 samples in each test and rising to 96 combined specimens when testing plasma fractionation. Pooled testing for COVID-19 could be combined with nucleic acid testing, to screen blood donations and reduce the risk of transfusion transmitted infections, with improved accuracy.

Methods using NGS are highly sensitive to the identification of microbes, even in pooled samples.^18^ To date, a total of 226 SARS-CoV-2 (severe acute respiratory syndrome coronavirus 2) sequences have been submitted to the National Center for Biotechnology Information (NCBI) GeneBank.^19^ NGS enables the identification of different strains present in the pool accounting for the contribution of different individual samples, which is not possible with RT-PCR. In the case of SARS, individual tracheal aspirate, a high-risk aerosol generating procedure, had the highest diagnostic yield by RT-PCR, followed by stool samples. Pooled throat and nasal swabs, rectal swab, nasal swab, throat swab, and nasopharyngeal aspirate specimens provided only a moderate yield. Procedures for the collection of stool and pooled nasal and throat swab specimens were the least hazardous, and the combination gave the highest yield for coronavirus detection by RT-PCR.^20^ For community surveillance of respiratory pathogens among adults and children, self-collected gargle samples have been found to be comparable or even more sensitive than throat swab samples with lower detectable Ct values in RT-PCR^21, 22^ and less bio-hazardous for collection by community healthcare workers.

At present, the World Health Organization (WHO) and the Indian Council of Medical Research (ICMR) recommend RT-PCR based tests for SARS-CoV-2 diagnosis as this test produces results within a few hours, is available in many countries on both manual and automated platforms, and can identify infected individuals from early infectious stages to recovery. Tests using viral antigens can detect COVID-19 three days after the onset of symptoms, however, in many countries these are not available. Rapid diagnostic tests using antibodies are unable to identify early cases as antibodies are only detectable 6-12 days after patients become symptomatic. In addition, tests using antibodies are not able to distinguish recovered cases from those who are currently infectious, however they may be useful in surveillance for individuals who have been infected with COVID-19.

The first pooled RT-PCR method suggested here for classifying the infection rate in a population using group testing is viable when population prevalence is low. When prevalence is high, then it is likely that the optimal size of the pool will have to be very small and the method is unlikely to present a dramatic reduction in resource use. On the other hand, we recommend using the second RT-PCR pooling method proposed to identify individuals only on samples from individuals who exhibit symptoms. The optimal pool size depends on the sensitivity of the test, and this is a function of viral load. Salivary viral load has been found to be highest in the first week after the onset of symptoms.^23^ While patients are asymptomatic, early in infection, the viral load is low making this technique unlikely to be useful in these individuals.

One limitation of pooling multiple RT-PCR samples is that the sensitivity of testing is reduced. To address this, it has been suggested that the number of samples being pooled be kept as low as possible to reduce dilution^24,25^ and special kits for extracting DNA from larger volumes be used.^8^ However, in some settings, pursuing optimal pool size may still not be possible due to fixed testing resources.

Herein we recommend the use of the pooled sample method with a binary hierarchical testing strategy for the detection of SARS-CoV-2 by RT-PCR in community surveillance particularly in resource limited settings where testing kits, facilities, and personnel are scarce. Our best estimates are that for the currently estimated prevalence of COVID-19 in many low- and middle-income countries, a single cluster of 30 tests could be combined from patients exhibiting cough, fever and other mild-flu like symptoms. Clustering should be undertaken at a geographic level to prioritize neighborhoods or localities for additional containment efforts.

## Data Availability

N/A

## References

1 Dong E, Du H, Gardner L. An interactive web-based dashboard to track COVID-19 in real time. The Lancet Infectious Diseases 2020; 0. DOI:10.1016/S1473-3099(20)30120-1.

2 CDC COVID-19 Response Team. Severe Outcomes Among Patients with Coronavirus Disease 2019 (COVID-19) — United States, February 12–March 16, 2020. MMWR Morb Mortal Wkly Rep 2020; 69. DOI:10.15585/mmwr.mm6912e2.

3 Imperial College COVID-19 Response Team. Impact of non-pharmaceutical interventions (NPIs) to reduce COVID-19 mortality and healthcare demand. 2020 https://doi.org/10.25561/77482.

4 Interim Guidelines for Collecting, Handling, and Testing Clinical Specimens from Persons for Coronavirus Disease 2019 (COVID-19). Coronavirus Disease 2019 (COVID-19). 2020; Published online Feb 11. https://www.cdc.gov/coronavirus/2019-ncov/lab/guidelines-clinical-specimens.html (Accessed March 26, 2020).

5 Emmanuel JC, Bassett MT, Smith HJ, Jacobs JA. Pooling of sera for human immunodeficiency virus (HIV) testing: an economical method for use in developing countries. J Clin Pathol 1988; 41: 582–5.

6 Currie MJ, McNiven M, Yee T, Schiemer U, Bowden FJ. Pooling of clinical specimens prior to testing for Chlamydia trachomatis by PCR is accurate and cost saving. J Clin Microbiol 2004; 42: 4866–7.

7 Van TT, Miller J, Warshauer DM, et al. Pooling nasopharyngeal/throat swab specimens to increase testing capacity for influenza viruses by PCR. J Clin Microbiol 2012; 50: 891–6.

8 Edouard S, Prudent E, Gautret P, Memish ZA, Raoult D. Cost-effective pooling of DNA from nasopharyngeal swab samples for large-scale detection of bacteria by real-time PCR. J Clin Microbiol 2015; 53: 1002–4.

9 Taylor SM, Juliano JJ, Trottman PA, et al. High-throughput pooling and real-time PCR-based strategy for malaria detection. J Clin Microbiol 2010; 48: 512–9.

10 Frosth S, König U, Nyman A-K, Aspán A. Sample pooling for real-time PCR detection and virulence determination of the footrot pathogen Dichelobacter nodosus. Vet Res Commun 2017; 41: 189–93.

11 Cortey M, Napp S, Alba A, et al. Theoretical and experimental approaches to estimate the usefulness of pooled serum samples for the diagnosis of postweaning multisystemic wasting syndrome. J Vet Diagn Invest 2011; 23: 233–40.

12 Schmidt GV, Mellerup A, Christiansen LE, Ståhl M, Olsen JE, Angen Ø. Sampling and Pooling Methods for Capturing Herd Level Antibiotic Resistance in Swine Feces using qPCR and CFU Approaches. PLoS ONE 2015; 10: e0131672.

13 Wu Z, Yang L, Ren X, et al. Deciphering the bat virome catalog to better understand the ecological diversity of bat viruses and the bat origin of emerging infectious diseases. ISME J 2016; 10: 609–20.

14 Du D, Hwang FK, Hwang F. Combinatorial group testing and its applications. World Scientific, 2000.

15 Knuth DE. Optimum binary search trees. Acta Informatica 1971; 1: 14–25.

16 Pooling Method for Accelerated Testing of COVID-19 | Technion - Israel Institute of Technology. https://www.technion.ac.il/en/2020/03/pooling-method-for-accelerated-testing-of-covid-19/ (Accessed March 26, 2020).

17 Stone A. Nebraska Public Health Lab begins pool testing COVID-19 samples. KETV. 2020; Published online March 26. https://www.ketv.com/article/nebraska-public-health-lab-begins-pool-testing-covid-19-samples/31934880 (Accessed March 26, 2020).

18 Next Generation Sequencing of Pooled Samples: Guideline for Variants’ Filtering. https://www.ncbi.nlm.nih.gov/pmc/articles/PMC5037392/ (Accessed March 27, 2020).

19 SARS-CoV-2 (Severe acute respiratory syndrome coronavirus 2) Sequences. https://www.ncbi.nlm.nih.gov/genbank/sars-cov-2-seqs/#nucleotide-sequences (Accessed March 27, 2020).

20 Laboratory Diagnosis of SARS. https://www.ncbi.nlm.nih.gov/pmc/articles/PMC3323215/ (Accessed March 27, 2020).

21 Comparison of gargle samples and throat swab samples for the detection of respiratory pathogens - ScienceDirect. https://www.sciencedirect.com/science/article/abs/pii/S0166093417300836 (Accessed March 27, 2020).

22 Detection of respiratory viruses in gargle specimens of healthy children - ScienceDirect. https://www.sciencedirect.com/science/article/pii/S1386653215000189 (Accessed March 27, 2020).

23 To KK-W, Tsang OT-Y, Leung W-S, et al. Temporal profiles of viral load in posterior oropharyngeal saliva samples and serum antibody responses during infection by SARS-CoV-2: an observational cohort study. The Lancet Infectious Diseases 2020; 0. DOI:10.1016/S1473-3099(20)30196-1.

24 Westreich DJ, Hudgens MG, Fiscus SA, Pilcher CD. Optimizing screening for acute human immunodeficiency virus infection with pooled nucleic acid amplification tests. J Clin Microbiol 2008; 46: 1785–92.

25 Muniesa A, Ferreira C, Fuertes H, Halaihel N, de Blas I. Estimation of the relative sensitivity of qPCR analysis using pooled samples. PLoS ONE 2014; 9: e93491.

